# Prospective Blinded evaluation of Thermalytix, an artificial intelligence-enhanced breast thermal imaging software, correlated with radiologist-interpreted mammograms: Results of an exploratory study in Zambia

**DOI:** 10.1101/2025.01.12.25320093

**Authors:** Maurice Mwale, Mutinta S. Nteeni, Peter Mwaba, Mercy N. Chipamp

**Author notes:** Corresponding author:* Maurice Mwale, University of the Witwatersrand, Johannesburg, South Africa. Competing interests: All authors declare no financial or non-financial competing interests.

## Abstract

**Background:** While mammography is commonly used for breast cancer detection, its widespread implementation in resource-constrained nations is challenging. Artificial intelligence-based Thermalytix is a low-cost, portable, radiation-free, automated test for breast cancer detection in women of all ages. Although used in India, the efficacy of Thermalytix has not been tested in an African population.

**Objectives:** To assess the agreement and correlation coefficient of Thermalytix output with radiologist-reported mammography, in a Zambian tertiary care population.

**Methodology:** In October 2023, 169 women were evaluated with both Thermalytix and standard mammography at Maina Soko Military Hospital at Lusaka. Thermalytix uses advanced machine learning algorithms to interpret breast thermal scans and generates a quantitative score indicating the likelihood of malignancy. All women underwent both tests, with results blinded both ways. Subsequently the Spearman correlation coefficient and level of agreement between Thermalytix output and BIRADs scoring from radiologist-interpreted mammography was calculated.

**Results:** 144 women with complete data were analysed in this report, with median age of 50 years (53.5% postmenopausal, 65.3% asymptomatic). Six women were assessed as mammography test positive and 138 as mammography negative; in these, the correlation between Thermalytix and mammography using Spearman test of rank correlation was 0.9 [very strong], and using the US FDA recommended test of agreement, positive agreement was obtained in 83.3%.

**Conclusion:** Demonstrating a very strong correlation and level of agreement with mammography, along with its good sensitivity, specificity and negative predictive value in previous clinical trials, Thermalytix has the potential to be an additional tool in the early detection of breast cancer in Zambia.

## 1. Introduction

Globally, breast cancer (BC) is the leading cause of cancer-related mortality among women (1). In Zambia, BC is the second leading cause of cancer mortality after cervical cancer, with an estimated incidence rate in 2022 of 20.0 per 100,000 women (2) and with observed mortality of 9.5 per 100,000 women (3). Thus, over 50% of women diagnosed with BC in Zambia will succumb to the disease. The principal factor driving this high disease-attributable mortality is the late stage at diagnosis prevalent in Zambia, reflecting delays in detection, diagnosis and treatment (4). Sixty percent of Zambian participants enrolled in the African Breast Cancer Disparities and Outcomes study (5) presented with late-stage disease, a trend consistent across various countries in Africa and other developing countries.

The World Health Organization (WHO) recommends population-based mammographic screening only in high-resource settings with well-funded, coordinated health systems (6). In low and middle-income countries (LMICs), the absence of sustainable financing, cultural barriers, and limited availability of medical equipment and healthcare workers [HCW] make systematic mammographic screening challenging to implement. For instance in the entire country of Zambia (with 10 million women population) there are only 16 mammography machines, 10 in public hospitals and 6 in private hospitals (7), with most located in second-level and third-level referral hospitals, thus excluding the majority rural population. Equally relevant is the limited availability of trained radiologists that results in mammography reporting times of up to 1 month (8). Further, in Africa, as in other emerging LMICs, BC occurs at a relatively younger age, when breast density is higher (9), further reducing the efficacy of mammographic screening. There are also socio-cultural barriers to screening (10) in Africa, such as fear of screening test results, lack of knowledge about the disease and screening, distance to the screening clinic, embarrassment to undergo screening, lack of support or permission from husbands to undergo screening, and high cost of screening. Thus most of the 10 million eligible women in Zambia do not receive regular breast screening which currently consists of only opportunistic approaches with contact points such as government hospitals, private clinics, and mobile screening camps.

For countries with limited resources, the WHO suggests a combination of clinical history, clinical breast examination (CBE), and diagnostic breast ultrasound. (6). However, a recent systematic review by Ngan et al (11) reported that the efficacy of CBE for breast cancer screening remains uncertain, with no consistent evidence demonstrating its ability to down-stage disease, or reduce mortality rates. While a cluster-randomised trial in India that used intensive CBE training demonstrated a non-significant 15% reduction in breast cancer mortality (12), a recent Cochrane review reported by Kenyan researchers (13) found limited improvements in CBE detection rates even with training of HCWs in CBE. Also, screening rates using CBE have remained poor across African nations.

We believe that this is the rationale to study other effective and affordable approaches to breast cancer detection. Further, while announcing the Global Breast Cancer Initiative in the year 2021 (14), the WHO emphasised that any breast cancer screening program that incorporates community awareness and mobilization, primary care-based screening, enhanced referral, and tracking processes to facilitate timely diagnosis and treatment is likely to be feasible, and effective in LMIC countries. For screening to be appropriate, it must be acceptable, equitable, accessible, sustainable, and economically efficient for the target population. (15).

Thermalytix is one such breast screening tool attempting to bridge this gap in early detection of BC. It is a low-cost, non-invasive, portable, radiation-free, and artificial intelligence (AI) based test for the detection of breast cancer in women of all ages and densities (16). Thermalytix automatically generates a quantitative score, indicating the presence of a possible malignancy by using state of the art machine learning algorithms and innovative medically interpretable thermal and vasculature-based radiomic features that closely represent the actual tissue-level metabolic and vascular activities observed on thermal imaging (17).

The authors were interested in evaluating Thermalytix in view of the existing clinical evidence of Thermalytix:-

A. It has demonstrated promising results in hospital-based prospective efficacy trials conducted in India. In one study, Thermalytix showed 10% non-inferiority compared to mammography in symptomatic women (18). Another study reported sensitivity and specificity of 100% and 92.41%, respectively, for asymptomatic women, and 89.85% and 82.39%, respectively, for symptomatic women (19). Another study in women with diverse breast tissue composition, including both dense and fatty breasts reported an overall sensitivity of 95.2% and specificity of 88.6% (20).
B. Community based studies conducted in India have demonstrated its feasibility and efficacy for population level breast cancer screening (21,22) over 15096 women with a cancer detection rate of over 3 times that of similar studies in the region that used CBE. It also has shown excellent patient satisfaction rating (23) as Thermalytix is a contactless test done in complete privacy of the individual.
C. Thermalytix has European CE MDD Mark with the following indication-‘it is approved to be used for screening and early diagnosis of breast cancer in women above 18 years in an environment where patient care is provided by healthcare personnel. Women who are found to be suspicious for malignancy by Thermalytix® should be referred for a confirmatory diagnostic test. Primary diagnostic and patient management decisions are made by a qualified healthcare professional’.

This was the background with which the authors sought to test Thermalytix in an African population with the primary goal-of evaluating the correlation between the output of automated Thermalytix with radiologist reported mammography in women in Zambia. A strong correlation would then guide our decision towards adopting this screening technology to complement the current clinical pathway for breast cancer detection that exists in Zambia.

## 2. Materials and Methods

### 2.1 Study Protocol and Population

This study was conducted between 10th October 2023 and 27th October 2023 at Maina Soko Military Hospital in Lusaka, Zambia. Women between 30 to 80 years, both asymptomatic and symptomatic, who had been referred for standard mammography, were recruited to this study aimed at evaluating the correlation coefficient and level of agreement between the output of Thermalytix with mammography. An ethics waiver was obtained from the Lusaka Apex Medical University Bio-medical Research Ethics Committee (LAMUBREC). Written informed consent was obtained from all consenting participants. All procedures were performed in compliance with relevant local laws and institutional guidelines and were conducted in accordance with the Helsinki Declaration as revised in 2013.

All participants underwent both thermal imaging with Thermalytix analysis and mammography [Figure 1]. Thermal imaging was performed before mammography to avoid image interference from breast compression.

**Figure 1:**
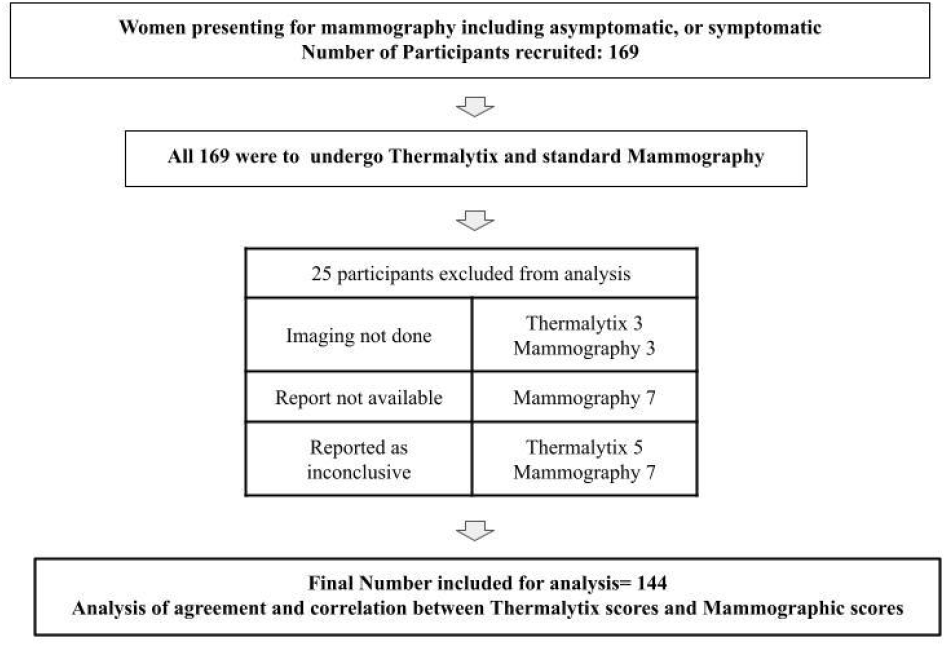
Schematic chart of the planned study

Imaging for the Thermalytix test was conducted by a hospital nurse after a formal two-day training program. The Thermalytix patient report was generated by the software automatically, as described later in section 2.3.

The mammogram was obtained by the hospital mammography technician who had completed a postgraduate diploma in mammography. The interpretation of mammography images were conducted by an experienced breast radiologist at the hospital.

Both the Thermalytix and mammography test results were kept blinded to each other until the time of analysis.

At this point, with participants recruited, the study concluded and analysis was performed. Further diagnostic imaging was guided by the treating physician following standard practice guidelines, and was not influenced by the Thermalytix output. This follow-up data was not available for this initial exploratory study.

### 2.3 Description of Thermalytix Screening

To perform thermal screening for Thermalytix test, infrastructure used included: an infrared camera [here, FLIR E54 thermal camera] with a spatial resolution of 320×240 and temperature sensitivity of 50 mK, a laptop connected to Thermalytix web-services, a portable air-cooler and an internet connection with minimum speed 2 MBPS. A private enclosure was created using curtains, so that the participant was neither seen nor touched at any time during the test [Figure 2A/2B].

**Figure 2:**
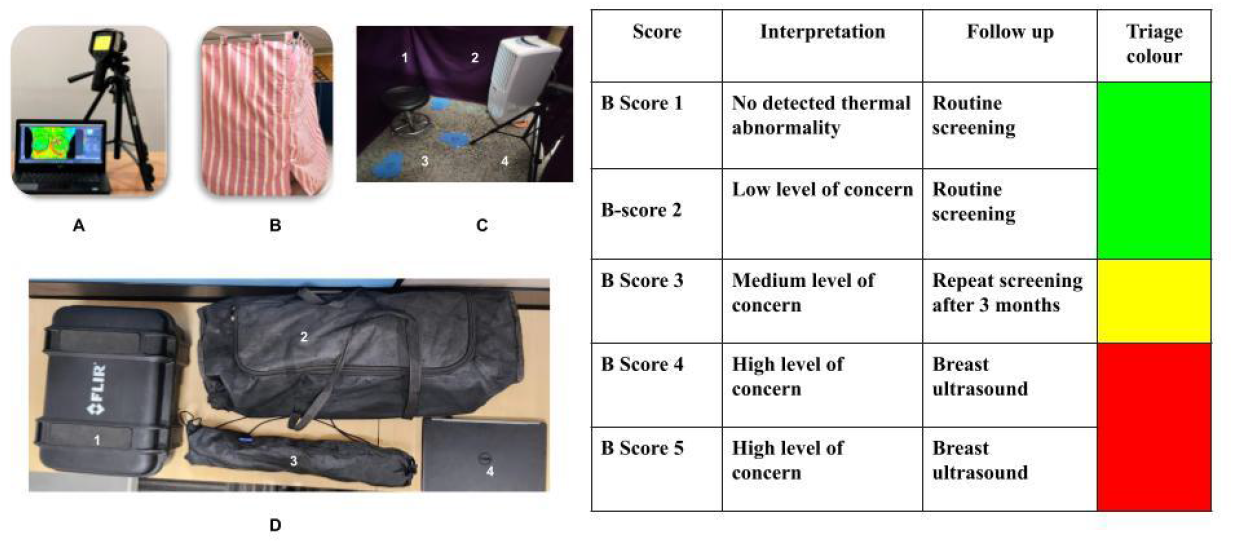
Left panel:-a) Thermalytix includes a portable thermal sensor, a laptop and cloud-based AI software. b) Screening is conducted inside an enclosure created with cloth curtains providing privacy. Throughout the entire process of screening, the technician is seated outside the enclosure. c) Within the both, are 1. a rotatable stool, 2. a portable air cooler, 3. foot markings to guide positioning of the women, 4. the thermal camers. d). The items needed to conduct screening may be folded in this manner for transportation-1. Camera case, 2. foldable screen structure with curtains, 3. Camera tripod, 4. Laptop. Right panel: depiction of the scoring system of the Thermalytix system.

As depicted in Figure 2C, the entire system is foldable allowing transport without a specialised vehicle. Setting up the screening merely involves creating the enclosure with collapsible aluminum rods and curtains, and setting up the camera, air-cooler and laptop and takes 10 minutes. As shown in Figure 2B, the screening is easily performed in any room that is available. The enclosure is 6 feet by 6 feet which ensures sufficient area for a patient to sit on a stool and rotate 180 degrees comfortably. The camera is placed 3 feet from the patient (sitting on the stool). The Thermalytix software runs on any standard android laptop. The camera operates via a standard electricity socket, and has battery back-up. The camera system requires no maintenance (only annual calibration).

The technician ensures that the participant is seated and is comfortable before leaving the screening area. The rest of the process is performed with the participant behind the closed curtains and the technician outside. From outside the enclosure, the technician asks the participant to remove their upper clothing to expose the body above the waist. The woman then sits in front of the infrared camera and air cooler for around 5 minutes. During this time, basic history related to breast disease, personal and family history of cancer is obtained, and chest wall cooling is monitored continuously by the technician using the software.

When thermal equilibrium is achieved, the tool prompts the technician to facilitate the capture of optimal data. The technician then directs the woman to use foot markings on the ground to obtain proper orientation for imaging and five thermal images are obtained, from the neck to the abdomen-frontal, left-oblique, left-lateral, right-oblique, and right-lateral. These images are uploaded to the cloud-based Thermalytix software, where pre-trained AI algorithms extract salient vascular and thermal radiomics features. These radiomic features are analysed by Thermalytix machine learning classifiers to generate instant patient reports with quantitative scores [Figure 2D]. An overall score called B-Score, analogous to BIRADS, indicates the likelihood of malignancy. To ensure ease of comprehension for all educational levels of HCW, a colour-coded Thermalytix output is also generated with a scale depicted in Figure 2D (for more information of the scoring system, please refer Bansal et al (21). Women categorised as test positive are advised to undergo further confirmatory diagnostic work-up. The entire process starting from patient preparation to report generation takes approximately 12-15 minutes per person.

### 2.3 Scoring methodology

For mammography, as per general clinical practice, test positivity was considered to ACR BIRADS 4 and 5 as per the radiologist’s report, while test negativity included BIRADS 1, 2, and 3. BI-RADS 0 were deemed inconclusive and excluded from the analysis.

The test positivity of Thermalytix was defined as B-scores of 3, 4, and 5, while test negativity included B-scores of 1 and 2. Cases with a B-score of 0 were classified as inconclusive and excluded from the analysis.

### 2.4 Statistical analysis

Data analysis was performed using SPSS (IBM SPSS v26.0.0.0). The extent of correlation between Thermalytix B-Score and mammographic BIRADS was performed by two tests: Spearman’s Correlation Coefficient(*ϱ*) which is a test of rank correlation measuring the strength of association between two ranked variables and US FDA recommended test of agreement between a new test and a non-reference standard [24]. In the absence of a true reference standard, measures like sensitivity and specificity cannot be calculated.

### 2.5 Funding

This research did not receive any specific grant from funding agencies in the public, commercial, or not-for-profit sectors

## 3. Results

Between October 10, 2023, to October 27, 2023, 163 women were recruited at Maina Soko Military Hospital in Lusaka, Zambia to evaluate the correlation of Thermalytix and mammography. Of these, 25 participants (15.3%) were excluded from the final analysis due to the following reasons [Figure 1]: in 6 women, either Thermalytix or mammography was not performed; in 7 cases, mammography reports were unavailable; in 7 cases, mammographic scoring was inconclusive; and in 5 women, Thermalytix scoring was inconclusive.

### 3.1 Demographics of study population

Among the 144 women who were considered for the analysis, the median age was 50 years (IQR 11 years), with 51.3% aged less than 50 years. The majority were postmenopausal (53.5%) and asymptomatic (65.3%). Demographic data are presented in Table 1.

**Table 1:** Demographics of the women screened.

| Variable | Number Screened [%] | Thermalytix Positive [%] | Mammography Positive [%] |
| --- | --- | --- | --- |
| **Age** |  |  |  |
| **<30** | 1 [0.7%] | 0 | 0 |
| **30-40** | 15 [10.4%] | 5 [33.3%] | 2 [13.3%] |
| **41-50** | 58 [40.3%] | 6 [10.3%] | 0 |
| **51-60** | 44 [30.6%] | 6 [13.6%] | 1 [2.3%] |
| **61-70** | 18 [12.5%] | 2 [11.1%] | 0 |
| **>71** | 8 [5.5%] | 7 [87.5%] | 3 [37.5%] |
| **Total** | **144** | **26 [18.0%]** | **6 [4.2%]** |
| **Menopausal Status** |  |  |  |
| **Pre -Menopause** | 67 [46.5%] | 12 [17.9%] | 2 [3.0%] |
| **Post -Menopause** | 77 [53.5%] | 14 [18.2%] | 4 [5.2%] |
| **Symptomatic Status** |  |  |  |
| **Asymptomatic** | 94 [65.3%] | 12 [12.8%] | 2 [2.1%] |
| **Symptomatic** |  |  |  |
| **Suspicion of breast lump (inclusive of other symptoms)** | 11 [22%] | 9 [81.8%] | 4 [36.4%] |
| **Breast Pain Only** | 21 [42.0%] | 3 [14.3%] | 0 |
| **Nipple Discharge Only** | 9 [18.0%] | 2 [22.2%] | 0 |
| **Possible Skin changes Only** | 1 [2.0%] | 0 | 0 |
| **Post-Mastectomy Asymptomatic** | 3 [6.0%] | 0 | 0 |
| **Post-Mastectomy: Symptomatic** | 1 [2.0%] | 0 | 0 |
| **Post-Lumpectomy: Asymptomatic** | 2 [4.0%] | 0 | 0 |
| **Post-Lumpectomy Symptomatic** | 2 [4.0%] | 0 | 0 |
| **Symptomatic** | 50 [34.7%] | 14 [28.0%] | 4 [8.0%] |

### 3.2 Correlation between Thermalytix B Scores and Mammographic BIRADS Scores

The Spearman coefficient (*ϱ*) was 0.90, indicating a very strong positive correlation between the scores generated by Thermalytix and those obtained by mammography [Table 3], while using the US-FDA recommended test of agreement, positive agreement between the scores were obtained in 83.3% [Table 3]. Calculations are presented in the Supplementary file.

**Table 2:**
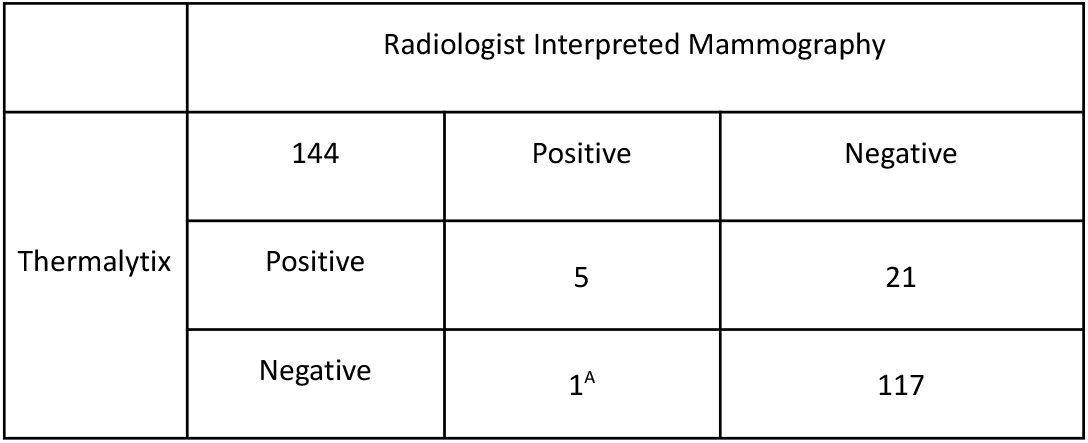
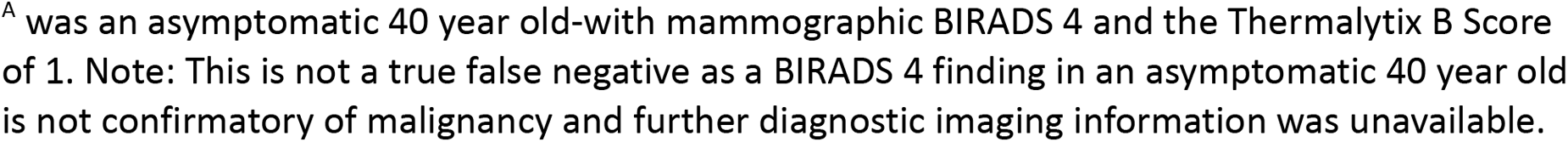
Contingency Table of the outputs of Thermalytix and Mammography.

|  | Radiologist Interpreted Mammography |  |  |
| --- | --- | --- | --- |
|  | 144 | Positive | Negative |
| Thermalytix | Positive | 5 | 21 |
|  | Negative | 1<sup>A</sup> | 117 |

<sup>A</sup> was an asymptomatic 40 year old-with mammographic BIRADS 4 and the Thermalytix B Score of 1. Note: This is not a true false negative as a BIRADS 4 finding in an asymptomatic 40 year old is not confirmatory of malignancy and further diagnostic imaging information was unavailable.

**Table 3:** Interpretation of Correlation and Agreement Scores.

| Spearman Rho |  |  |  |
| --- | --- | --- | --- |
| Interpretation | Very Strong | Greater than 0.7 | Spearmann<br>Correlation of<br>Thermalytix was<br>0.90 |
|  | Strong | 0.40 to 0.69 |  |
|  | Moderate | 0.3 to 0.39 |  |
|  | Weak | 0.2 to 0.29 |  |
|  | Absent | 0 to 0.19 |  |
| Test of agreement |  |  |  |
| Measure | Percentage | Low limit | High limit |
| Positive Percent Agreement | 83.3% | 43.7% | 97.0% |
| Negative Percent Agreement | 84.8% | 77.9% | 89.8% |
| Overall Percent Agreement | 84.7% | 78.0% | 89.7% |

## D. Discussion

While Thermalytix had been earlier evaluated and compared with standard radiological imaging in an Asian population, this is the first experience of the usage of Thermalytix in an African population. In this study conducted in Zambia, Thermalytix demonstrated strong to very strong positive correlation and 83.3% positive agreement with radiologist-reported mammography with radiologist-reported mammography.

This result presents a promise to enable equitable care. In the African continent, around 85.2% of countries belong to LMICs economies, where most residents rely on underfunded public health care systems, and have limited access to clinical expertise, or to secondary or tertiary health centers. Therefore, affordable AI based tools may prove beneficial in triaging patients at the primary care level itself before they present to secondary or tertiary care centres, thereby improving the efficiency of specialists even in secondary care and easing the case burden caused by a worsening health provider-to-patient ratio (25).

Although CBE as a screening measure is being practised as a national program in Zambia and in other African countries, we feel that the bulk of the literature on CBE published thus far only highlights its many limitations. Ngan et al (11) has reported an overview of systematic reviews of CBE, including 11 reviews, and reported that population coverage rates in screening programs based on CBE are suboptimal and the efficacy of CBE in down-staging disease or in reducing mortality remains unproven. This is likely due to a combination of factors

1. The performance of CBE is entirely dependent on the skill of the examiner, with no standardisation of technique, and no guidelines for optimal reporting (26). Since the most sensitive technique has not been established, published clinical results report variable sensitivity of CBE of 40% to 69%, with a pooled sensitivity of 54·1% (11).
2. To be useful, effective CBE requires a trained and diligent community HCW whose competence is trusted by the community. However, in most of Africa, the HCW has many roles, is overburdened (27) and in the face of competing healthcare priorities does not prioritise cancer screening. Of note, a recent Cochrane review found limited improvements in CBE detection rates even with imparting of training to HCWs in CBE, again implying a need to strengthen any CBE-based screening service (13). Since Thermalytix is an computer-based automated test, there is no variability in analysis with minimal dependency on the skill of the technician.
3. Sociocultural barriers represent the most significant barrier to breast screening in most studies (10) in both high-income and LMICs and are not lessened by CBE. Cultural norms of modesty such as embarrassment, shame on disrobing, husband’s disapproval, and certain religious beliefs prevent women from accessing services requiring undressing in front of an HCW. Since Thermalytix is a completely privacy conscious test where no one sees or touches the woman during the test, this socio-cultural barrier is being addressed by the Thermalytix test.

We believe that Thermalytix would be useful as an additional tool towards the early detection of breast cancer especially in situations where mammography is unavailable. We anticipate benefits for public health utilisation as the system is low cost, lightweight, portable, and requires minimal physical infrastructure and thus may be feasible in diverse healthcare settings.

Further it is non-invasive, non-contact, radiation-free, and does not require breast compression, making the user experience highly acceptable for patients. The AI-based automation of Thermalytix with its easy-to-use user interface allows the entire process from image capture to report generation to be performed even by a semi-skilled HCW at the point of care. The software mandates automated image quality checks thus ensuring standardised image capture. As compared to CBE which is a subjective and qualitative assessment, the use of Thermalytix also allows objective and quantitative analysis of any abnormality detected, improving the chance of improved cancer detection rates.

In the last decade, developments in radiomics-based AI algorithms have been applied to breast thermal images, enabling interpretation of the minute temperature variations not with the human eye but with computer-aided thermal image interpretation (17). These machine learning algorithms may eliminate the subjectivity of the manual interpretation process through automated quantitative analysis, wherein the interpretation process becomes more objective, more quantitative and score based. This approach is being explored at various centres across the world for screening and detection of breast cancer, especially where other imaging modalities, like mammography are not universally available (28).

The recent advancement of AI into primary health may be a transformative event with potential to mitigate healthcare disparities (29). These systems improve primary healthcare in a variety of areas, including by improving risk prediction, supporting clinical decision making, increasing the accuracy and timeliness of diagnosis, optimising resource allocation and reducing work-burden of the primary healthcare workforce. Of note, technologies utilising AI for image analysis generally alleviates shortages in skilled personnel and affordable systems and address the constraints of availability of medical equipment. Already, AI algorithms are in use for interpreting medical images in X-rays, MRIs, and CT scans, early detection of diabetic retinopathy and diagnostic imaging of the cervix (30).

We acknowledge certain limitations of the study. Firstly, the sample population was small, and hence may not be representative of all the subsets of women encountered in Zambia. Secondly, this study was designed to compare the correlation and agreement between Thermalytix and mammography; in the absence of a true reference standard we are unable to comment on the sensitivity and specificity for Thermalytix from this small study

## 5. Conclusion

This study demonstrates the feasibility of using Thermalytix in an ethnically African population in Zambia and demonstrates very strong positive correlation, and good positive agreement between Thermalytix and mammography. This initial experience has prompted us to plan for a larger study using Thermalytix towards widespread adoption for breast cancer screening in Zambia.

## Data Availability

All data produced in the present work are contained in the manuscript

## Acknowledgment

We thank Niramai Health Analytix, Bangalore, India for providing the thermal camera and Thermalytix tool for this study.

## Ethics Approval

An ethics waiver has been obtained for this study from the Lusaka Apex Medical University Bio-medical Research Ethics Committee (LAMUBREC).

## Supplement A: Calculation of Spearman’s Coefficient Calculation

Supplement A: Calculation of Spearman's Coefficient Calculation
|  | Thermalytix<br>Score | Mammography<br>Score | Rank 1 | Rank 2 | d<sub>i</sub> |
| --- | --- | --- | --- | --- | --- |
| 1 | 77 | 112 | 5 | 5 | 0 |
| 2 | 42 | 23 | 4 | 4 | 0 |
| 3 | 5 | 3 | 1 | 2 | 1 |
| 4 | 12 | 5 | 3 | 3 | 0 |
| 5 | 9 | 2 | 2 | 1 | 1 |

The sum of the differences in ranks squared 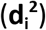.

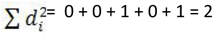

The following formula was then used 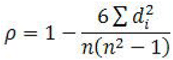

Substitute the sum of 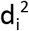 information and the number of scores variable (*n*) into the formula:

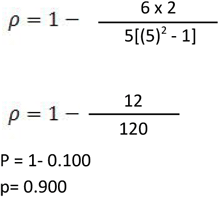

Therefore, the Spearman coefficient is **0.900**.

[Statistics without maths using SPSS for Windows. Authors: Christine P. Dancey, John Reidy … Print Book, English, 2004. Edition: 3rd ed ISBN, 013124941X, 9780131249417]

## Supplement B: Test of Agreement between a new test and a non-reference standard

|  | Comparative method |  |  |  |
| --- | --- | --- | --- | --- |
| Test method |  | Positive | Negative | Total |
|  | Positive | 5 | 21 | 26 |
|  | Negative | 1 | 117 | 118 |
|  | Total | 6 | 138 | 144 |

| Summary statistics |  |  |  |
| --- | --- | --- | --- |
| Measure | Percentage | Low limit | High limit |
| Positive Agreement PPA | 83.3% | 43.7% | 97.0% |
| Negative Agreement PNA | 84.8% | 77.9% | 89.8% |
| Overall Agreement POA | 84.7% | 78.0% | 89.7% |

https://tools.westgard.com/two-by-two-contingency.shtml [see reference 24]

